# Understanding preconception care practices, beliefs, and attitudes in Australian primary care: A qualitative study of health professionals

**DOI:** 10.1101/2025.01.16.25320103

**Authors:** Cherie Caut, Danielle Schoenaker, Erica McIntyre, Amie Steel

## Abstract

**Background:** Primary care is well placed to provide preconception care. To support primary care professionals in meeting the preconception care needs of prospective parents, an understanding of their perspectives and experiences of providing preconception care is needed. As women consult with a range of primary care health professionals for preconception care, including general practitioners (GPs), midwives, and naturopaths, this study aimed to explore these health professions’ beliefs and attitudes towards preconception care and describe their preconception care practice behaviours in Australia.

**Methods:** Qualitative methods were employed. Focus groups and interviews with health professionals (n=18) in clinical practice (more than 5 years) within Australia were conducted between May and August 2021. Health professionals comprised GPs (n=6), midwives (n=5), and naturopaths (n=7) and were recruited through professional organisations. Fieldwork explored the practice services provided, beliefs and attitudes towards preconception care, and preconception care practice behaviours. Data analysis used a framework thematic analysis approach.

**Results:** Three major themes were identified: *Defining preconception health and care* (subthemes: *defining preconception health* and *defining preconception care*)*, Understanding primary practitioners’ role in preconception care* (subthemes: *the GP’s role as central to preconception care*, *role is holistic, educational, and empowering* and *role of personal experience and gender in being a preconception care provider*), and *Situating preconception care in primary care practice* (subthemes: *preconception care patient populations*, *preconception care within broader health services* and *preconception health information sources are varied*).

**Conclusions:** While health professionals shared similar views on the factors that comprise preconception health, some components of preconception health differed among the health professional groups. Although it is universally agreed that GPs are key providers of preconception care, they are not the only health professionals with a role. A wider range of health professionals could aid in meeting the preconception care needs of people of reproductive age with greater coordination among them. To improve the provision of multi-disciplinary preconception care further insights into shared and complementary responsibilities among health professionals in primary care settings are needed.

## Background

Preconception care aims to address preconception risks among prospective parents to improve maternal and child health outcomes [1, 2]; for example, through intervention strategies to improve nutrition and health behaviours before conception [3, 4]. Multiple preconception risks and health behaviours are modifiable (i.e., capable of being changed), including body composition (e.g., underweight, overweight, obesity), lifestyle behaviours (e.g., alcohol, caffeine, smoking, physical activity), nutrition (e.g., folic acid, multivitamins), environmental exposures (e.g., radiation, pesticides, chemicals), and birth spacing (e.g., short interpregnancy intervals), and can affect maternal and child health outcomes [5, 6]. Some of the maternal health outcomes associated with modifiable preconception risks include preeclampsia, gestational diabetes mellitus and antenatal and postnatal depression, among others [5]. Some of the child health outcomes associated with modifiable preconception risks include preterm birth, low birth weight, macrosomia, congenital abnormalities, and neurocognitive disorders [5, 6]. Preconception risks also include teenage pregnancy, advanced maternal age, mental health conditions (e.g., depression, bipolar disorder), intimate partner violence, infectious diseases (e.g., sexually transmitted infections, immunisations status), genetic disorders (e.g., cystic fibrosis, fragile X, thalassemia), chronic diseases (e.g., diabetes, epilepsy), and medication use [7].

Primary care settings are well placed with opportunities to provide preconception risk screening and preconception health education and interventions [8, 9] to prospective parents. Women consult various health professionals for preconception care, including general practitioners (GPs) and midwives[8]. GPs have contact with the general population [9] with opportunities to screen prospective parents for preconception risks and view their role as the primary providers responsible for preconception care [10]. Midwives have also been found to agree that preconception health promotion is part of their role and are well placed to promote preconception health to women planning a pregnancy in the future [11, 12]. Having contact with women between pregnancies [11, 12], midwives can provide interconception care in primary care settings [9, 13]. Naturopaths are also primary care providers in Australia who treat various health conditions [14], with health promotion as a core principle of naturopathic practice [15, 16]. Naturopaths have opportunities to promote preconception health information during the preconception period; contemporary evidence suggests women attempting to conceive are more likely to consult with a naturopath than women who are not attempting pregnancy [17] and seek assistance from naturopaths for preconception care [18].

Evidence suggests that some health professionals may not always be aware of their potential role in preconception health promotion [8, 19] and that it is unclear which groups of health professionals are responsible for delivering preconception care [20, 21]. Some primary care health professionals, including GPs and midwives, regard reproductive health care as the responsibility of obstetricians and gynaecologists and family planning or sexual health clinicians [22]. These health professionals may also perceive that women are going elsewhere for their reproductive health care or that they will provide it only if the woman asks [22]. An understanding of primary care health professionals’ (i.e., GPs, midwives, and naturopaths) current perspectives and experiences of providing preconception care in clinical practice settings across Australia will help to inform preconception care policy and health service planning to support health professionals in meeting the preconception care needs of prospective parents.

### Objective

This research aimed to explore the beliefs and attitudes towards preconception care and describe the preconception care practices and the preconception information-seeking behaviour of general practitioners, midwives, and naturopaths in Australia.

## Methods

### Research design

A phenomenological theoretical framework underpinned the study design [23] and was chosen as an appropriate methodology for the research objectives. Qualitative methods were employed to facilitate an exploration of complex phenomena encountered by health professionals providing preconception care [23]. Focus groups and semi-structured interviews were utilised as data collection methods [23].

### Participants

Between May and August 2021, GPs, midwives, and naturopaths in current clinical practice for at least five years in Australia were invited to attend an online focus group or interview.

### Setting

Focus groups and interviews were conducted online with GPs, midwives, and naturopaths. Facilitating the focus groups and interviews online allowed participants to attend from various geographical locations within Australia, including remote regions. It was a particularly important setting for facilitation during the COVID-19 pandemic. The Zoom Video Communications platform was used to maintain face-to-face group discussions and ensure the interpersonal interaction central to focus group and interview methods was maintained in the online environment. This setting also allowed for audio recordings to occur at the time and be transcribed later.

Furthermore, online settings are beneficial for exploring participants’ perspectives on topics that may be sensitive [24]. Focus groups were facilitated by two members of the research team (AS, CC). Interviews were conducted by one member (CC) of the research team.

### Recruitment

Participants were recruited through direct email, newsletter, and social media channels through professional organisations: Royal Australian College of General Practitioners (RACGP), RACGP Specific Interest group Antenatal and Postnatal Care, Australian College of Midwives (ACM), Naturopaths and Herbalists Association of Australia (NHAA), and Complementary Medicines Association (CMA). To compensate for their time, a gift voucher was offered to each health professional (AUD $50.00 for midwives and naturopaths; AUD $100.00 for GPs). This amount was selected based on input from stakeholder groups (RACGP), published data about practitioner hourly fees in clinical practice [18] and ensuring equality between groups [25]. A short screening survey was developed in Qualtrics XM™ online survey software [26] for interested health professionals to view the participant information sheet and eligibility criteria and provide their consent to participate in the study. The screening survey asked questions to obtain respondents’ preferred availability for organising the focus groups and interviews and contact information for communication with researchers CC and AS. Purposive sampling methods were used to select participants [23]. After determining the most suitable date and time for participants, an invitation to participate in the relevant focus group was emailed. Figure 1 details the phases of recruitment and retention of participants in the study.

**Figure. 1.**
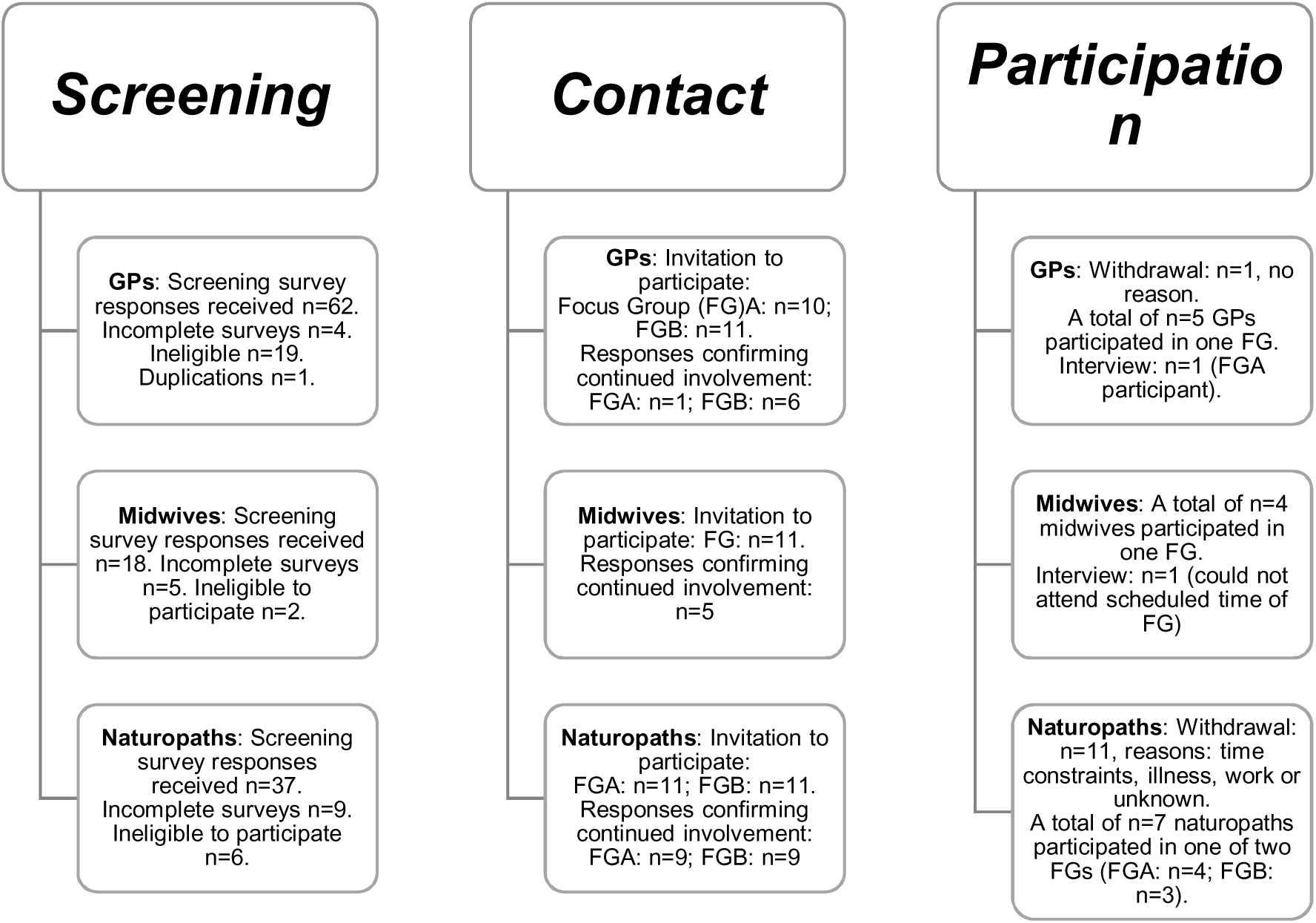
Participant recruitment and retention for focus groups and interviews with health professionals.

### Sample

In total, 18 health professionals participated in either online focus groups or interviews comprising GPs (n=6), midwives (n=5), and naturopaths (n=7). One focus group and one-on-one interview each were held online for GPs and midwives, and two focus groups were held for naturopaths. Participants in each focus group ranged between three to five (GPs: [n=1 focus group; n=5 participants], midwives: [n=1 focus group; n=4 participants], naturopaths [n=2; focus group A, n=4 participants; focus group B, n=3 participants]). Each focus group was between 80 and 90 minutes, and the interviews were 30 to 40 minutes.

### Data collection

Participants were asked to share their perspectives and experiences as health professionals providing preconception care. A focus group and interview guide (see Additional file 1) were developed to facilitate the focus group discussion, guide the interview and explore three domains: practice services provided, beliefs and attitudes towards preconception care, and preconception care practice behaviours.

### Data analysis

Two research team members (CC, AS) analysed data from transcripts using a framework thematic analysis approach [27]. This approach is an established analysis method within health service research employing five stages: familiarisation, identifying a thematic framework, indexing, charting, and mapping and interpretation [27]. Data from the focus groups and interview audio recordings were transcribed by Rev audio transcription service [28] and imported into NVivo 12 Pro qualitative data analysis program software [29]. No new themes were identified relative to the research objectives of sixteen focus group participants; the interviews with one GP and midwife each confirmed this, as no new themes were identified. The data results were descriptively reported using the ‘Consolidated criteria for reporting qualitative research (COREQ): a 32-item checklist for interviews and focus groups [23]. Quotes were selected based on the quality and the theme’s representativeness.

## Results

Thematic analysis of the focus group and interview data identified some shared experiences and perspectives of providing preconception care in the Australian healthcare setting across all or some health professions, while other experiences were unique to one health profession. Three major themes were identified: *defining preconception health and care*, *understanding primary practitioners’ role in preconception care*, and *situating preconception care in primary care practice*.

### Defining preconception health and care

The first theme covered participants’ views and understanding of how to define preconception health and care. These concepts were defined differently, with one subtheme addressing ‘defining preconception health’ and the second subtheme ‘defining preconception care’.

### Preconception health is multifactorial, and for both biological parents

Participants shared their views on the meaning of the term ‘preconception health.’ Components of preconception health identified by different participants across all focus groups encompassed the health of the person or couple, substances and medications use and other modifiable preconception risk factors. One dominant definition shared by naturopaths and midwives was the belief that preconception health is the overall health of the parent, whether they are single or in a couple:

> *“Preconception health is their health. The health of the couple preparing to conceive, or the single woman, or the donor.” (Naturopath 1)*

> *“Obviously, it takes two people to make a baby…And make sure they [men] are just as healthy as the women.” (Midwife 3)*

As seen above, this naturopath participant specifically mentioned that preconception health is important for anyone to conceive or donate gametes (i.e., ova, sperm), and this is inclusive of single women and men. This more expansive view, which highlighted that any person contributing to genetics to the conception of a child requires attending to their preconception health, was also shared by another naturopath:

> *“The foundational aspects of both whomever the people are who are going to be providing the genetic material for the baby, what their levels of health are.” (Naturopath 7)*

Several naturopaths also expressed the view that preconception health is difficult to define as it is evolving, multifactorial and complex, as outlined by this naturopath:

> *“Preconception health is constantly evolving. I don’t think we can put a definition on it as such, and if we even tried to, we’d have to be talking for hours because there’s so much involved.” (Naturopath 1)*

In addition to the broad concepts outlined in the above quotes, participants also identified specific components that require attention during preconception care. The preconception health components identified from descriptions within each health professional group are summarised in Table 1.

**Table 1.**
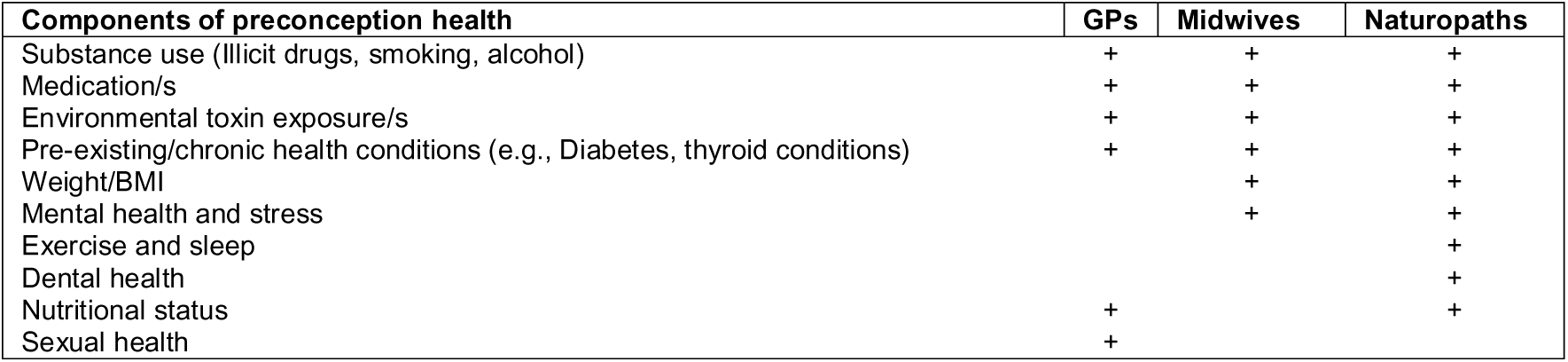
Components of preconception health by health profession.

Participants from each health professional group shared that the aspects important to consider for preconception health were substance use (e.g., illicit drugs, smoking, alcohol), medications and environmental exposures, as described within these quotes from representatives across the different health professional groups:

> *I would go through their medications. Again, it’s focusing on if any of those could be teratogenic in that pregnancy. I would be checking first smoking, alcohol, drugs…and also…occupation, because if they’re being around radiation or something like that.” (GP 6)*

> *“A lot of people are using a lot of chemicals and they may not realise that they can be teratogens for the early pregnancy. (Midwife 5)*

> *“It is a lot of environmental toxins…I think the importance of all of that role in the preconception health.” (Naturopath 4)*

Pre-existing or chronic health conditions, such as diabetes and thyroid conditions, were identified as important to preconception health by participants, as described in these quotes:

> *“You’re looking for things like diabetes, thyroid problems.” (GP 6)*

> *“I think women that have medical conditions, then that’s a real trigger for them…they live in this reality of having diabetes, for instance, how will a pregnancy affect diabetes?” (Midwife 5)*

> *“I actually get a lot of people who have difficulty falling pregnant, and their hormones are imbalanced, and their thyroid is imbalanced.” (Naturopath 6)*

Also considered as important to preconception health by midwife and naturopath participants was ideally being at a healthy weight before pregnancy:

> *“Looking at their physical wellbeing, is the mother at a healthy weight? Or would it be beneficial for her to get a healthier body weight?” (Midwife 5)*

> *And really, they have to have got healthy prior to that. So, lost the weight prior to that. I mean, this is in the ideal scenario, of course. (Naturopath 4)*

Midwives and naturopath participants also identified mental health as important to preconception health:

> *“Looking at their mental health, whether they need any support around that.” (Midwife 5)*

> *I’m always referring for mental healthcare plans as well a lot of the time, so definitely some sort of psychological support. (Naturopath 3)*

Further to mental health as an important aspect to consider for preconception health, several naturopaths individually described stress as a factor, as captured by this quote:

> *“The stress factor I’ve noticed plays a big role over the years.” (Naturopath 6)*

GPs shared the view, along with naturopath participants, that nutritional status and risk of nutrient deficiencies were important considerations for preconception health:

> *“If they’re vegetarian, they may need some supplements.” (GP 6)*

> *“And I think the things that get ignored so much now, like…general nutritional depletion and all that sort of stuff.” (Naturopath 7)*

Uniquely, naturopath participants described that lifestyle health practices such as exercise and sleep were important to preconception health, as captured in this quote by a naturopath:

> *“You need to have to some extent a balanced lifestyle in terms of exercise, enough sleep.” (Naturopath 6)*

Naturopath participants were also the only group to identify dental care as part of preconception care, as this naturopath described:

> *“I also get them to go off and to see their dentist because having dental work done whilst pregnant isn’t such a good idea, so I always send them off to their dentist if they haven’t been in the last couple of months.” (Naturopath 5)*

An aspect of preconception health distinctively identified by GP participants was sexual health and sexually transmitted infections as being an important consideration for preconception health, as captured by these GPs’ quotes:

> *“Sexual history, to look at [the] risk of STIs.” (GP 6) “Sexual [health] STI’s, those sorts of things.” (GP 4)*

### Preconception care involves screening, counselling and education

The concept of ‘preconception care’ identified through this theme primarily included concepts such as screening tests, providing counselling, and educating future parents. As demonstrated by the following quotes from a naturopath, GP, and midwife, each type of health professional mentioned screening blood tests as primarily related to identifying vitamin and mineral deficiencies, including some specifically mentioned nutrients including iron, vitamin B12, and folic acid across all health professions and vitamin D was included by naturopaths and midwives as captured among these quotes across the different health professionals:

*We do their bloods…a lot of our women are very low in iron…we get all those basic bloods done” (Midwife 4)*

> *“They’re all without fail vitamin D deficient, iron deficient. Often have not had folate.” (Midwife 3)*

> *“Whether we need to supplement, whether there’s low iron or low B12 and also to start the folic acid.” (GP 2)*

> *“That you’ve got the right amount of vitamin D and the right amount of iron, and your active B12 is in, and your folate is all at optimal levels.” (Naturopath 5)*

GPs and midwife participants, but not naturopaths, specifically stated that cervical screening tests (or ‘CST’) as a part of the prenatal screening tests comprised preconception care:

> *Making sure they’ve had the pap smears or CST, as we call it now, during the first consult. It’s part of my investigation. (GP 1)*

> *And then in terms of the testings that we would do, so we’ll check if their pap smear is up to date…all patients planning a pregnancy, we order a group of blood tests. (GP 6)*

> *“We do this cervical screening; we do their bloods.” (Midwife 4)*

Only GP participants described screening for genetic conditions, sexually transmitted infections, and vaccination status as captured in these quotes by two GPs:

> *“I think that genetics, from that perspective, for them understanding what options they have for genetic testing.” (GP 1)*

> *I really tend to focus on the vaccinations and the STI screens when I’m doing my preconception. (GP 2)*

Each type of health professional shared that providing education regarding preconception health risks was a part of preconception care. However, the specific types of risks described as examples among them varied. Educational counselling, as described by the health professional groups, included managing expectations related to family planning regarding birth spacing, maternal age and the potential impact of these preconception risks regarding fertility outcomes. Midwife and GP participants described including education on birth spacing, as seen in these quotes by a midwife and GP who highlighted the importance of discussing previous births and future birth plans:

> *“It’s also really important to debrief and talk about their previous birth…and counselling around that before they consider conceiving again.” (Midwife 2)*

> *“What their actual family planning, how many kids they want, what space of time and how old they are and understanding from that perspective” (GP 1)*

In comparison, this naturopath participant described the importance of education on the timing of conception:

> *“Educational and timing conception…and the importance of that.” (Naturopath 4)*

There was a general agreement between each type of health professional that preconception care included counselling for education on food safety practices, dietary nutrition, and physical activity as described in these quotes:

> *“Educating them on foods they should avoid or other bits and pieces from that.” (GP 1)*

> *“Education around diet and exercise…changing what they’re eating and their portion sizes and that sort of thing.” (Midwife 5)*

> *“Encourage them to do as much of the lifestyle, and the diet…and then taking their supplements.” (Naturopath 1)*

Also shared among participants was the view that providing education on supplements is important for a healthy pregnancy and that addressing nutritional deficiencies is a part of preconception care. These quotes provide examples of the various nutrients described as examples across participants from each health profession, including folic acid, vitamin B12, vitamin D, and iron. GPs were the only health professional group to describe iodine in their examples:

> *“And make sure that you’re [the patient] on folic acid and iodine.” (GP 4)*

> *“Often have not had folate, or any sort of preconception supplement or anything like that beforehand.” (Midwife 3)*

Naturopaths were the only health professional group that described fish oil containing high DHA and antioxidants in their examples:

> *“I choose the supplement according to their history…the B-vitamins…high DHA fish oil…the antioxidants…vitamin D…iron…active B12…folate is all at optimal levels.” (Naturopath 5)*

Naturopath participants described the unique experience that preconception care was also considered in the context of optimising natural fertility and potentially improving assisted fertility outcomes. This naturopath emphasised the important role of being healthy to become pregnant:

> “If somebody comes in and they’re sitting in front of me and they say oh, I’m desperate to have a child, I just say, “You need to get healthy, when you get healthy and when your everything is in sync, then you will fall pregnant naturally.” (Naturopath 6)

This naturopath highlighted the role of antioxidants on egg quality as being an aspect to address:

> *“Look at the antioxidants in their diet and usually top up with antioxidants to protect egg quality.” (Naturopath 5)*

Also specifically related to improving egg quality, this naturopath described women seeking preconception care before assisted reproductive cycles to improve the outcome:

> *“You still get many…women who are doing IVF, who are like yeah, I’m starting an IVF cycle next week, and I want to improve my egg quality.” (Naturopath 7)*

### Understanding primary practitioners’ role in preconception care

The second theme identified covered participants’ views and understanding of primary practitioners’ roles in preconception care. One subtheme addressed ‘the GP’s role as central to preconception care,’ and the second addressed the ‘role is holistic, educational, and empowering.’ The third subtheme addressed the ‘role of personal experience and gender in being a preconception care provider.’

### GP’s role is central to preconception care

Across health professions, the belief that preconception care fits directly within the role and responsibility of GPs was consistent. Participants justified this belief by the view that GPs are the first contact for health care and, therefore, they should be the primary provider of preconception care, as suggested by these quotes from a midwife and naturopath participants:

> *“I think GPs are sort of the gatekeepers of people’s first point of call whether they have the time or the knowledge around that.” (Midwife 5)*

> *“The GP is the first port of call for many patients.” (Naturopath 4)*

GPs shared the view that they are trusted to provide health care within the general community and are usually the first point of contact for health care for their patients:

> *“GPs are synonymous with the general public, as trustworthy people, they are your first point of call.” (GP 1)*

While participants from all health profession groups held this view of GPs’ importance in preconception care, naturopath and midwife participants asserted that they also hold valuable roles as primary care providers of preconception care.

Regarding preconception care, one naturopath described their role as synonymous with a GP’s role:

> *“I think naturopaths are uniquely placed to do this work, and we do offer something that pulls everything together, I think. Like the GP, we can pull everything together.” (Naturopath 4)*

Midwife participants felt they had a valuable role to play as one midwife describes they have more time to spend providing effective preconception care in comparison to GPs:

> *“A lot of preconception care is about education and empowering women to know, and that requires time, and that’s the value of the midwives because the GPs have got seven minutes or whatever they’ve got, and that’s the thing.” (Midwife 2)*

### Role is holistic, educational, and empowering

Another major shared belief expressed by participants was how they viewed their role as health professionals within the community. Participants perceived their role as being holistic health carers, GPs and midwives, specifically used the term ‘holistic’ in their descriptions:

> *“Preconception health and care are very holistic, and I think that’s what GPs do, we’re holistic carers.” (GP 1)*

> *“I think it’s holistic care. It’s in every aspect of their life. You’re always wanting to help and educate, and empower people to make better healthy choices, whether they’re wanting to have a baby or not.” (Midwife 2)*

This naturopath participant defined the holistic concept by describing that they follow the principle of treating the whole person:

> *“It’s that basic principle of treating the whole person…we look at that person and we go, we take a full case, we take a full health history and family history.” (Naturopath 7)*

Participants from all health professions perceive their role as being educators to their patients concerning preconception health information as exampled by this GP:

> *“Diet, supplements, foods, educating them on foods they should avoid or other bits and pieces from that.” (GP 1)*

Naturopath and midwife participants described how the responsibility to educate extended beyond their patients to include providing education to other health professionals in formal education settings or through the publication of authored materials and presenting health information to other health professionals as well as the public as described in these quotes from a midwife and naturopath:

> *“I talk to the educators, and I talk to them about how we do this.” (Midwife 2)*

> *“I’ve presented the naturopathic community with a lot of information through books and seminars over the years and trained a number of health professionals.” (Naturopath 1)*

Each type of health profession group described the views that their role was to empower their patients to make health behaviour changes through the education and counselling they provide as captured in these descriptions by a GP and midwife:

> *“I think the place that I can make the most difference, personally, is often the opportunistic stuff where I can identify someone who is usually at risk of an unplanned pregnancy that may want to continue, and I can see things that would really be important to the health of that pregnancy.” (GP 5)*

> *“So, I think preconception care was really trying to empower women to make choices that are good for them.” (Midwife 1)*

One naturopath participant specifically described how improving a parent’s health literacy has the potential to improve not only their health but also the health of their children as seen in this quote:

> *“I think that’s another important part of it, is that we’re putting them back out into the world as more health literate and more empowered to look after themselves and their health, which therefore makes them more likely to look after their children well as well.” (Naturopaths 7)*

Midwives and naturopaths both described an aspect of their role as involving counselling as suggested in these quotes:

> *“I think midwives and nurses counsel all the time.” (Midwife 2) “At that point, we go into my counseling side of things, we do some counseling.” (Naturopath 6)*

One naturopath further described this aspect of their role as specifically comprising motivational counselling, particularly related to lifestyle health behaviour change, as captured in this quote:

> *“And I think the other part of our role that I see is motivational. They go to the doctors and they’ve got a BMI of 30 plus, and the doctor just tells them to lose a bit of weight and they don’t tell them whether it’s good to lose it quickly or slowly. We know the damage that can be done to the eggs when they’re losing weight. There’s all of those issues. But how do they drop that weight? It’s not like they haven’t been trying, perhaps. So, I think our role is motivational with lifestyle change with some people that’s bigger than others.” (Naturopath 4)*

### Role of personal experience and gender in being a preconception care provider

Across all health professions, participants described that being female or through their own experiences of pregnancy was another way they believed they had acquired some of their knowledge of preconception health and preconception care as exampled in these quotes from a midwife and naturopath:

> *“But otherwise, I guess, through my own pregnancies” (Midwife 5) “Some of it from being a woman myself” (Naturopath 6)*

One GP participant described how they believed there was an assumption that being female meant you knew preconception health information for providing preconception care as captured in this quote by a GP:

> *“People just assumed being a female I would have all the answers to the contraception and pre-counseling needs and preconception needs.” (GP 2)*

### Situating preconception care in primary care practice

The third theme identified covered participants’ views and understanding of situating preconception care in primary care practice. One subtheme addressed ‘preconception care patient populations,’ and the second addressed ‘preconception care within broader health services.’ The third subtheme addressed the ‘preconception health information sources are varied.’

### Preconception care patient populations

Participants described the patient populations seen by each type of health professional and these are reported in Table 2. Common across all health professions were adult women of reproductive age.

**Table 2.** Patient populations seen by each type of health professional.

| Population(s) | GPs | Midwives | Naturopaths |
| --- | --- | --- | --- |
| Adult women of reproductive age | + | + | + |
| Low socio-economic populations | + |  |  |
| High socio-economic population | + |  |  |
| Families | + |  |  |
| Adolescent females | + |  |  |
| Women coming off their contraceptive |  | + |  |
| Women in high-risk settings for domestic violence |  | + |  |
| Women from Indigenous groups | + | + |  |
| Women wanting to improve fertility |  |  | + |
| Women egg donors |  |  | + |
| Pregnant women | + | + |  |
| Women in the antenatal and postpartum period | + | + |  |
| Women with reproductive health conditions | + |  | + |
| Women trying to conceive | + |  | + |
| Male partner of women trying to conceive | + |  | + |

GPs reported providing care to families and adolescents; both GPs and midwives specifically provided services to pregnant women. Midwives and GPs described providing services to Indigenous groups of whom English is not a first language, and GPs described both low and high-economic-status populations accessing their services as exampled in these quotes by a GP and midwife:

> *“We’re a mixed building and a really varied demographic from quite low socio-economic through to very well-off people, really very diverse…small Aboriginal population in Ballarat, so [I] have [a] few Aboriginal patients.” (GP 4)*

> *“I work in very remote Aboriginal communities in Central Australia…I work predominantly with women who for whom English is not their first language.” (Midwife 1)*

Naturopaths described a unique perception that women wanted to improve their fertility and women egg donors used their services. Both GPs and naturopaths shared the view that they provided their service to women with reproductive health conditions and women trying to conceive, including their male reproductive partner.

### Preconception care within broader health services

Participants described their services as health professionals extending beyond preconception care, and these are reported in Table 3. Across all health professions, participants described being providers of family planning services, preconception care, and women’s health care. Naturopath participants otherwise described themselves as providers of fertility support services as suggested in this quote by a naturopath:

> *“I have probably talked about myself more as a women’s health and fertility specialist” (Naturopath 7)*

**Table 3.** Type of health services provided by health professionals.

| Type(s) of health service | GPs | Midwives | Naturopaths |
| --- | --- | --- | --- |
| General health care | + | + | + |
| Private practice | + | + | + |
| Family planning | + | + | + |
| Preconception care | + | + | + |
| Women's health care | + | + | + |
| Sexual health care | + |  |  |
| Public women's hospital |  | + |  |
| Birth classes |  | + |  |
| Remote community midwife |  | + |  |
| Emergency births |  | + |  |
| Multidisciplinary/integrated practice |  |  | + |
| Fertility support |  |  | + |
| Gastrointestinal health care |  |  | + |
| Pregnancy, antenatal and postpartum care | + | + |  |

Whereas GP and midwife participants described themselves as pregnancy, antenatal, and postpartum care providers as captured in these quotes from a midwife and GP:

> *“And I provide antenatal, labor and birth, and postnatal care in the woman’s home. And that continues from when she books in with me until six weeks after the baby’s born.” (Midwife 5)*

> *“From preconception times to family planning to pregnant and antenatal care, postpartum care and then when their kids come through doing the immunizations and the post-partum care from that perspective and it’s quite a broad spectrum.” (GP 1)*

GP participants distinctively described themselves also as a provider of gynecological and sexual health care, as seen in these quotes by GPs:

> *“When I have people coming with any gynecological problems. So if they’ve got period problems or any conditions around that, then that’s usually a discussion that will come up because sometimes, those things can affect fertility.” (GP 6)*

> *“That’s one of the things I provide as well, preconception as well as sexual health advice.” (GP 2)*

### Preconception health information sources are varied

Participants all reported using handouts to provide their patients with preconception health information. The most dominant way participants reported acquiring the information they knew regarding preconception health and preconception care was through professional development courses and seminars. Colleagues, professional training, and clinical experience were other avenues described by participants for having acquired knowledge on preconception health and care. GP and midwives’ participants also described accessing online information (e.g., health department websites) as a resource for preconception health information to share with their patients as captured in these descriptions by a GP and midwife:

> *“I actually always give them the Raising Children’s website, the government website, I think that’s a really great resource for them to start reading about more the diet stuff, because a lot of them want to ask the ‘Do’s and Don’ts’” (GP 5)*

> *“Also had some links for reputable websites, and I’ve used that.” (Midwife 1)*

Meanwhile, midwives and naturopaths reported using a naturopath-authored book on natural fertility as a source of preconception health information, as described in these quotes from a midwife and naturopath:

> *“Reading Natural Fertility by Francesca Naish and those sorts of books.” (Midwife 5)*

> *“We used Francesca’s book and we have over the years.” (Naturopath 4)*

Naturopath participants were unique in providing their experiences of accessing preconception health information from product companies, describing the importance of trust in that the information is accurate, as captured in this quote from a naturopath:

> *“So, we do have to trust that this information that we’re getting through our industry is correct, and I think most of us do.” (Naturopath 1)*

## Discussion

This study provides key insights into the experiences and perceptions of three groups of primary care health professionals providing preconception care in clinical practice settings within Australia: GPs, midwives and naturopaths.

While the participants in this study shared similar views on the factors that comprise preconception health and their care practices, some components were only described by one or two health professional groups. These variations may be due to health professionals approaching preconception health and care through the lens of their training or clinical experiences. For example, GPs are providers of general medical care, including health promotion and disease prevention education, and their inclusion of sexual health care (e.g., STI screening), genetic carrier screening, and vaccinations as a part of preconception care is synonymous with their medical training, skills, and general scope of practice [30]. Meanwhile, naturopaths’ consideration of exercise, sleep, and dental health as part of preconception health is also synonymous with their practice. Naturopaths are holistic healthcare providers that promote preventive and lifestyle health behaviours, which are foundational to their practice [15, 16, 18].

Another explanation for variations in the type of preconception health and care considerations by the type of health professional could relate to opportunistic patient contact. For example, in this study, both midwives and GPs specifically described educating their patients on birth spacing and emphasised the importance of discussing previous births and future birth plans with their patients. Midwives and GPs, by nature of their roles (e.g., GPs being providers of general health care including postnatal care and midwives providers of perinatal care) [30, 31] have contact with women throughout their reproductive years. The interconception period provides the unique opportunity to ask the One Key Question^®^ (OKQ^®^)[22] regarding women’s plans for future pregnancies and discuss the importance of birth spacing in that window [11–13]. The OKQ^®^ is a pregnancy desire screening question designed for clinicians in primary care (e.g., GPs and midwives) to identify reproductive health information and services women need for contraception, preconception, or interconception care [22] and could explain the experiences of the GPs and midwives related to education around birth spacing and family planning.

Health professional groups may cover differing aspects of preconception health and the preconception care they provide, as these findings suggest. Further insights into shared and complementary responsibility among health professionals in primary care settings are needed to improve the provision of multi-disciplinary preconception care.

Our study findings highlight that it is universally agreed that GPs are the central providers of preconception care; however, they are not the only health professionals involved in this care. Our findings suggest that a range of potential health professionals who have contact with women and men of reproductive age could also have a role in providing preconception care and that any health professional who has an established and trusting relationship could contribute to an individual’s preconception care. These findings align with other studies and recommendations from preconception care clinical practice guidelines [32–34]. While preconception care is best suited for primary care, and GPs are the main providers, other health professionals have a responsibility to contribute to providing preconception care, screen women for their pregnancy intentions and refer to specialised health services when preconception risks and medical conditions are beyond the skills and ability of the health professional [32–34].

The findings in this study suggest that each health profession provides only aspects of preconception care. While the participants in this study described several healthcare services and populations they provided for, differences were noted across the health professional groups. These findings highlight that while independent health professionals may provide aspects of preconception care, they may not be able to meet all the preconception care needs for the community, particularly for women concerning reproductive health outcomes. An integrated healthcare model could consider utilising all health professionals capable of addressing the reproductive health needs of the community across the life course [18, 34].

Coordinating care among health professionals in primary care is an acknowledged healthcare issue[35, 36], and given the lack of clarity among health professionals who have the responsibility to provide preconception care [20, 21] and poor communication between health professionals [21], resolving this issue is challenging. An integrated approach between health professionals who can deliver preconception care is needed and requires healthcare policies that address primary care coordination [35, 36]. Recently, Hall et al. proposed a model of community-based preconception care that comprises family planning, contraception, and preconception care provided by a range of trained health professionals [34]. Future research attention should be given to exploring preconception care health outcomes when preconception care services are provided within healthcare settings.

The findings from this study highlight that midwives and naturopaths could play an important role in preconception health promotion aimed at increasing the preconception health literacy of professional and public communities. Although each type of health profession in our study views their role as an educator to their patients, naturopath and midwife participants believed their educational responsibilities extended beyond their patients to include other health professionals and the public. The primary care provider aims to engage and empower individuals and communities to improve health outcomes [37], and health literacy is a critical predictor of empowerment [38]. Therefore, health education is a central tool for improving the health literacy of a population [38]. The role and scope of contemporary midwifery practice in Australia include promoting public health and empowering relationships to support the health and well-being of women and their families throughout pregnancy, birth, and the antenatal and postnatal period [39].

Similarly, for naturopaths, health promotion and patient education [16] are among the core principles of naturopathic practice, and most naturopaths (98%) engage in community education and health promotion activities [16]. The educational activities provided by naturopaths may provide effective preconception healthcare solutions within communities as they provide patient-centred care to motivated groups [16]. Future research that explores the capacity (e.g., skills, time) of midwives and naturopaths to provide professional and public preconception health education in communities is needed.

Our study reveals a potential gender bias towards women in preconception care. All the health professional participants in our study were women. Being a woman was viewed by participants as being one source of preconception health knowledge through their own lived experiences. Furthermore, the participants in our study described how they perceived that other healthcare providers assumed that being a woman meant they knew about preconception health and care. Gender bias towards women in preconception care does exist [40], and until recently, there has been little focus on men’s preconception health and reproductive outcomes[6]. One explanation for this is that there has been an over-emphasis on women in preconception care, as fertility has been viewed as a woman’s domain [41]. The lack of male focus in preconception care to date is evident, as shown in an Australian study where an overwhelming percentage (90%) of GPs (n=304) in the study providing care to men reported not feeling confident in their knowledge of modifiable factors that affect male fertility [42]. Men’s preconception health can have an impact on maternal and offspring health outcomes [6].There is a need for health professionals providing preconception care to be male-inclusive to encourage male involvement [41], and as suggested by the findings in our study, women exceed male health professionals providing preconception care. Future research, practice and policy attention is needed to address gender bias towards women in preconception care.

### Limitations and Future Directions

This study presents novel findings across three primary care professions involved in preconception care in Australia. However, it is not without limitations. Firstly, the smaller focus group sizes were at the minimum ideal for focus group dynamics [43, 44]. The study also only sampled Australian health professionals and as such may reflect characteristics specific to the Australian health system and context. Despite, these limitations the study still identifies new insights that warrant further exploration. In particular, due to the nature of qualitative research and the self-selection of patients, the data should not be seen as generalisable and should be further investigated through survey research to identify self-reported practice behaviours in the broader community of these health professions. Similarly, third-party observational studies can be employed to triangulate health professionals’ self-reported practice behaviours concerning preconception care.

### Conclusions

While the health professionals in this study shared similar views on the factors that comprise preconception health, some components of preconception health differed among the health professional groups. Likewise, health professionals shared similar views on what comprises their preconception care, with some differences noted among health professional groups for some preconception care practices. This could be because GPs, midwives and naturopaths consider preconception care from the lens of their own professional or clinical experiences. GPs are recognised by multiple health professions as central providers of preconception care; however, they are not the only health professionals with a role. Midwives’ and naturopaths’ capacity (e.g., skills, time) to provide professional and public preconception health education in communities must be further explored. While independent health professionals may provide aspects of preconception care, they may not be able to meet all the preconception care needs of the community. With greater coordination among health professionals, a wider range of healthcare service providers could aid in meeting the preconception care needs of people of reproductive age. Future research, practice and policy attention are needed to address gender bias among health professionals in preconception care. Future research focus should be given to exploring preconception care outcomes when preconception care services are provided within healthcare settings.

## Declarations

### Ethical approval and consent to participate

The research was approved by the University of Technology Sydney Human Research Ethics Committee (ETH20-5547).

### Consent for publication

Not applicable

### Availability of data and materials

The datasets used and/or analysed during the current study are available from the corresponding author upon reasonable request.

### Author contributions

The authors confirm their contribution to the paper: study conception and design: CC, AS; data collection: CC, AS; analysis and interpretation of results: CC, AS; draft manuscript preparation: CC, AS, DS, EM. All authors reviewed the results and approved the final version of the manuscript.

## Supporting information

Additional file 1. Focus group and interview guide

## Data Availability

All data produced in the present study are available upon reasonable request to the authors

## Acknowledgements

The authors thank member associations RACGP, RACGP Specific Interest group Antenatal and Postnatal Care, ACM, NHAA and CMA representing GPs, midwives, and naturopaths for supporting this research by promoting the study to their members for participation. The authors also thank each health professional who gave their time to participate in the study. The authors thank Endeavour College of Natural Health for supporting the research study by awarding a project grant.

## Additional files

Additional file 1. Focus group and interview guide

## List of abbreviations

ACM: Australian College of Midwives
CMA: Complementary Medicines Association
GP: General practitioner
NHAA: Naturopaths and Herbalists Association of Australia
RACGP: Royal Australian College of General Practitioners

