## Additional file 1. Focus group and interview guide for "Understanding preconception care practices, beliefs, and attitudes in Australian primary care: A qualitative study of health professionals"

Table 1. Focus group and interview guide

| **Domain** | **Questions** |
| --- | --- |
| ***1. Practice services provided*** | a. Tell me about how you see your role as a health professional within the community and the type of health services you provide? b. What populations does your health service provide for? |
| ***2. Beliefs and attitudes towards preconception care*** | a. What do you understand of the term preconception health? What do you understand of the term preconception care? b. How important do you perceive the provision of preconception health information and preconception care to be? c. How would you describe the role/responsibility of yourself and others in your profession to provide preconception health information, and preconception care? d. What other health professions, if any, do you feel should contribute to providing preconception health information and preconception care in the community? e. What do you think of the current level of focus on preconception health, and preconception care in the health-care system?   f. What do you believe to be the most appropriate timing/life stage for providing preconception health information, and preconception care? |
| ***3. Preconception care practice behaviours*** | a. An individual or couple come into your clinic and state that they want to start a family, how do you respond to that? b. What circumstances would prompt you to start a conversation with your patient/s regarding preconception health? Are there any circumstances in which you would not start a conversation….? c. What role do you expect male reproductive partners to play in the preconception period? How do you communicate this to women and their partners? d. Tell me about the type of preconception health information you provide to your patients. e. How confident are you in providing preconception health information? f. Why do you feel confident in providing this information? OR, if not confident, why do you think you lack confidence in providing this information? g. Is your confidence in providing information different for female and male patients? h. What resources have you accessed to find out the preconception health information you know? |
